# Early Diagnosis of Vascular Ehlers-Danlos Syndrome Through AI-Powered Facial Analysis: Results from the Montalcino Aortic Consortium

**DOI:** 10.1101/2024.10.20.24315773

**Authors:** David R. Murdock, Adarsh Suresh, Ernesto Calderon Martinez, Isabella Marin, Frances Marin, Alan C. Braverman, Angela T. Yetman, Shaine A. Morris, Dianna M. Milewicz

**Affiliations:** Department of Internal Medicine, University of Texas Health Science Center at Houston (UTHealth), Houston, Texas, USA; Cardiovascular Division, Department of Medicine, Washington University in St Louis, St Louis, Missouri, USA; Department of Pediatrics, Children’s Hospital & Medical Center of Omaha, Omaha, Nebraska, USA; Section of Pediatric Cardiology, Department of Pediatrics, Texas Children’s Hospital and Baylor College of Medicine, Houston, Texas, USA

## Abstract

**Purpose:** Vascular Ehlers-Danlos syndrome (vEDS), caused by *COL3A1* pathogenic variants, is a rare heritable aortic and arterial disorder associated with early mortality, mainly due to spontaneous vascular dissections and ruptures. Improved methods for diagnosing vEDS are needed so that guideline-based management can be initiated to prevent deadly complications and differentiate vEDS from overlapping conditions like hypermobile EDS (hEDS).

**Methods:** We implemented an AI facial recognition model based on the PhenoScore framework using a support vector machine (SVM) trained on facial images of thirty individuals, aged 6-65 years, with vEDS from the Montalcino Aortic Consortium (MAC), control images from the Chicago Face Database (CFD), and publicly available images of individuals with hEDS. Cross-validation was used to train the SVM, and statistical measures to evaluate the model performance were calculated. Local Interpretable Model-agnostic Explanations (LIME) was used to generate facial heatmaps highlighting the features driving the model’s predictions.

**Results:** The AI classifier showed excellent performance with as few as thirteen vEDS training images and distinguished vEDS from both controls and individuals with hEDS with high accuracy, achieving an area under the receiver operating characteristic curve (AUC) ≥ 0.97. LIME highlighted facial regions already established to characterize the facial features of vEDS patients (e.g., prominent eyes).

**Conclusion:** Our results demonstrate the potential of AI-based facial analysis for diagnosing vEDS. This method democratizes the early diagnosis of vEDS by reducing dependence on genetic testing, enabling optimal management and improved outcomes, particularly in resource-limited areas.

## Introduction

Vascular Ehlers-Danlos syndrome (vEDS) is a rare autosomal dominant genetic disorder caused by pathogenic variants in *COL3A1*, which encodes type III collagen^1^. The major complications of vEDS include arterial dissections and ruptures, and gastrointestinal (GI) perforations^2^, leading to a median survival of only 51 years^3^. Despite advancements in clinical management, approximately 70% of individuals with vEDS are only diagnosed after a major complication^3^, underscoring the urgent need for screening methods to identify these patients so that guideline-based management to prolong survival can be initiated^1,4^. Furthermore, vEDS needs to be differentiated from other conditions with overlapping systemic features, such as hypermobile Ehlers-Danlos syndrome (hEDS), a more common condition that does not predispose to life-threatening vascular or GI complications^5,6^.

Barriers exist that hinder rapid diagnosis even when vEDS is suspected, including the limited availability of genetic specialists^7,8^. Additionally, the majority of low- and middle-income countries (LMICs) have limited access to genetic testing^9,10^. Related conditions, such as Marfan syndrome, are diagnosed early based on prominent systemic features and aortic root enlargement on echocardiography. In contrast, vEDS cases lack dramatic systemic features and typically have normal echocardiograms. Given these challenges, alternative methods of identifying and confirming vEDS cases are needed. We sought to determine whether artificial intelligence (AI)- based facial analysis could identify individuals with vEDS with high sensitivity and specificity given vEDS-associated facial features that include a thin vermilion of the lips, a narrow nose, and prominent eyes^9,10^.

AI-based facial image analysis platforms like Face2Gene (FDNA Inc., USA) and GestaltMatcher have been used to diagnose rare genetic diseases^11,12^. These tools have successfully identified conditions such as 22q11.2 deletion syndrome^13^, Williams syndrome^13,14^, and Noonan syndrome^14,15^, though performance can vary widely depending on the specific gene involved, as with Cornelia de Lange syndrome (CdLS)^16^. Recently, AI-based facial image analysis has been shown to successfully identify individuals with Marfan syndrome with a high level of accuracy^17^. Here, we describe a highly accurate AI-based tool that is sensitive and specific for distinguishing vEDS cases from controls and overlapping conditions based on facial images.

## Materials and Methods

Facial images of individuals with vEDS were obtained from the Montalcino Aortic Consortium (MAC) patient registry, which contains clinical and genetic data from 1,680 individuals worldwide with pathogenic variants (PVs) and variants of uncertain significance (VUSs) in genes for heritable thoracic aortic disease (HTAD)^18^. Thirty images of patients with vEDS were used, including 20 from cases with glycine missense variants affecting the triple helical domain of type III collagen, seven with splice-site variants predicted to cause in-frame exon skipping, and three with frameshift, nonsense or full gene deletion variants leading to haploinsufficiency (HI). Additionally, 21 facial images from adult White females with hEDS were obtained from social media and other public sources. As a control dataset representative of the general population, we included images from the Chicago Face Database (CFD, version 3.0)^19^. The main CFD contains images of 597 male and female individuals between the ages of 17-65 recruited in the United States who self-identify as Asian, Black, Latino, or White. The baseline demographic characteristics of the studied image datasets are provided in Table 1.

**Table 1:** Baseline characteristics of image datasets analyzed in this study.

|  |  | vEDS | hEDS | CFD Controls |
| --- | --- | --- | --- | --- |
|  | Images, n | 30 | 21 | 597 |
| Demographics | Adults ($\geq 18$ years), n (%) | 24 (80) | 21 (100) | 594 (99.4) |
|  | Age (years), average (range) | 34.3 (6-65) | NA | 28.9 (17-65) |
|  | Female, n (%) | 23 (76.7) | 21 (100) | 307 (51.4) |
|  | Asian, n (%) | 1 (3.3) | 0 (0) | 109 (18.3) |
|  | Black, n (%) | 0 (0) | 0 (0) | 197 (33) |
|  | Latino, n (%) | 3 (10) | 0 (0) | 108 (18) |
|  | White, n (%) | 26 (86.7) | 21 (100) | 183 (30.6) |
| *COL3A1*<br>Variant<br>Type | Glycine | 20 (66.7) |  |  |
|  | Splicing | 7 (23.3) |  |  |
|  | HI | 3 (10) |  |  |

vEDS, vascular Ehlers-Danlos syndrome; hEDS, hypermobile EDS; CFD, Chicago Face Database; HI, haploinsufficiency, NA, data not available

For image analysis, we implemented an AI workflow adapted from the open-source PhenoScore framework^20^ (Fig. 1A). The initial step involved preprocessing images by detecting, cropping, and aligning faces to standardize the input for facial feature extraction which was then performed by VGGFace2^21^, an advanced convolutional neural network (CNN) algorithm trained on a diverse database of 3.3 million facial images, spanning various ethnicities, sexes, and ages. This process generated feature vector encodings that represented the facial characteristics identified by the neural network. These facial encodings, from both cases and controls, were then passed into a support vector machine (SVM) to train a classifier, where the number of cases and controls were matched to prevent class imbalance. As part of this process in PhenoScore, stratified 5-fold cross-validation was performed to partition the data into training and test subsets, ensuring balanced class distributions while evaluating performance across different data splits. This approach also incorporated hyperparameter C tuning to optimize model performance, minimize the risk of overfitting, and ensure consistent generalization across various data subsets. The PhenoScore framework’s primary metric used to assess the classifier’s ability to discriminate between cases and controls is the Brier score^22^, ranging from 0 to 1, with lower scores indicating better performance. Brier scores are useful for evaluating the accuracy of probabilistic predictions in machine learning^23^. In addition, an area under the receiving operator curve (AUC, higher is better) value reflective of the model’s overall performance was calculated.

**Figure 1:**
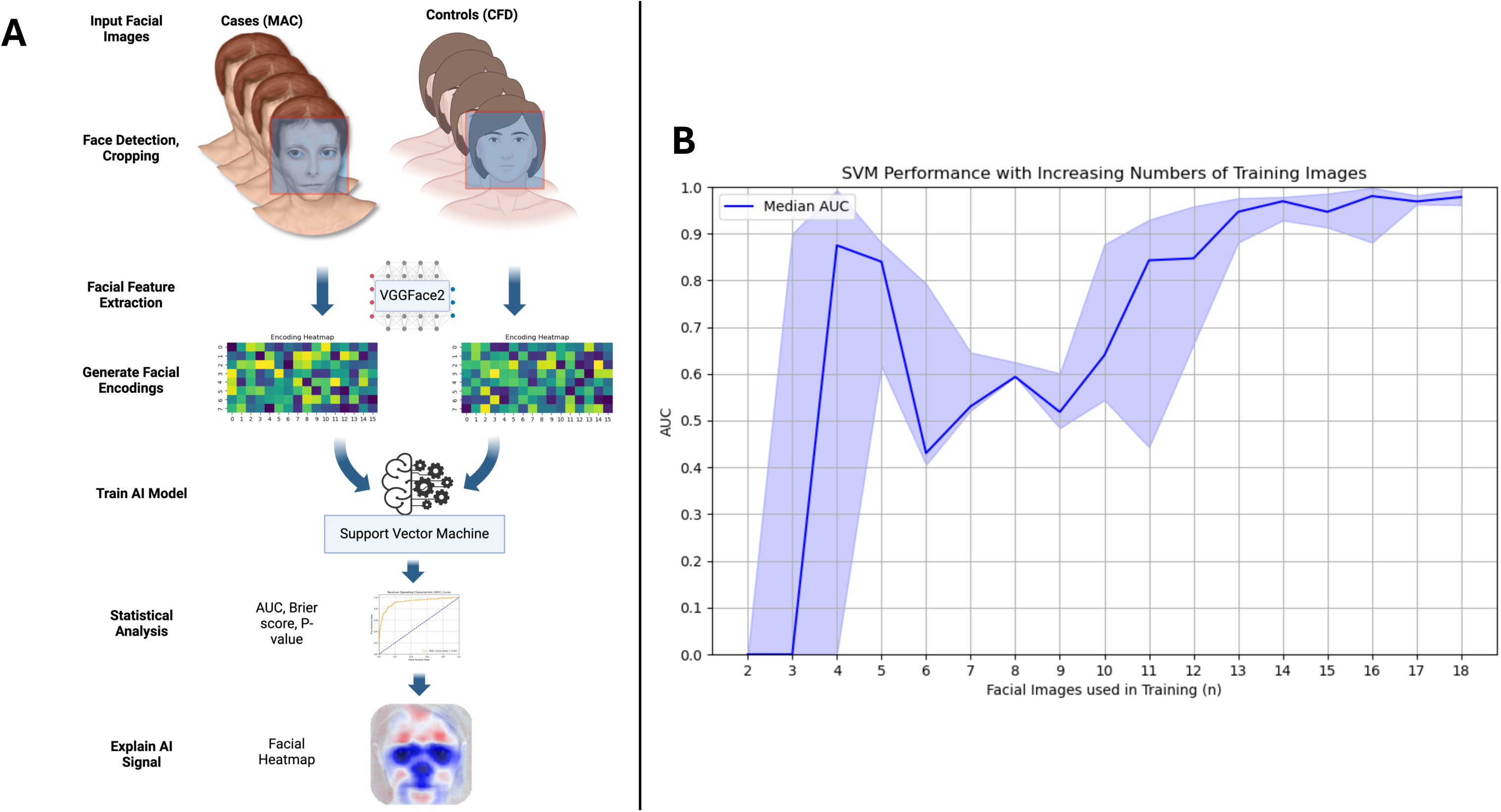
AI-Powered Facial Analysis for Diagnosis of vEDS. Panel A illustrates the workflow: image acquisition, facial feature extraction/encoding, SVM training, statistical analysis, and LIME heatmap generation. The heatmap background uses a StyleGAN-generated synthetic face^33^. Panel B depicts model performance with an increasing number of vEDS and matched control images used in the training set. Median AUC and 95% confidence intervals are plotted. Excellent predictive accuracy (Brier score of 0.13, AUC of 0.94 (95% CI 0.8894, 0.9899), p-value of 1.35×10^-6^) is achieved with thirteen images. vEDS, vascular Ehlers-Danlos syndrome; SVM, support vector machine; LIME, Local Interpretable Model-agnostic Explanations; and AUC, area under the receiver operating characteristic curve

To create a distribution of performance metrics under the null hypothesis (i.e., that the model’s performance is not significantly better than chance), the image labels were randomly shuffled, and the SVM was retrained on the shuffled data. After shuffling, the original Brier score was compared to the scores from the randomly permuted labels using a one-sided Mann-Whitney U test to generate a p-value, as implemented by PhenoScore. For comparisons against CFD controls, 100 training permutations were performed, and the entire process was repeated ten times using a different randomly selected set of controls. For analyses of variant type subgroups and vEDS vs. hEDS, 1000 permutations with ten repeats were used, as the higher number was necessary due to dataset imbalances and the inability to repeatedly sample controls. Finally, statistical metrics from each repeat iteration were either averaged or combined using Fisher’s method, as implemented in PhenoScore.

To determine the minimum sample size required to effectively train the model to distinguish between vEDS cases and controls, we initially focused on the largest subgroup within the MAC registry: 18 adult White females with vEDS. To create a comparable control group, we selected 18 age-, sex-, and ethnicity-matched facial images from the CFD. This careful matching ensured that any observed differences were primarily attributable to the syndrome rather than sex or ethnic/racial features. We trained the SVM incrementally, starting with two images from the vEDS and matched control groups. The training set size was increased by one image in each group until all 18 vEDS images were included, and model performance was recorded at each step to evaluate the minimum number of images needed to achieve an AUC ≥ 0.9.

We then trained the SVM on the full vEDS cohort and control images from the entire CFD dataset. Although the vEDS cohort was limited in diversity, the decision to use the full CFD dataset aimed to enhance generalizability across different ages, sexes, and ethnicities and reduce overfitting to specific demographic features. For analyses involving the hEDS cohort, sex-, age-, and race-matched CFD images were used, as the hEDS group only contained White females. Due to dataset size limitations, the *COL3A1* variant type analysis compared glycine substitution cases (n=20) to a combined group (n=10) of splice site and haploinsufficiency (HI) cases.

To highlight the facial features driving the model predictions, we used the Local Interpretable Model-agnostic Explanations (LIME) function within PhenoScore to generate facial heatmaps emphasizing the key distinguishing features^24^. LIME works by iteratively perturbing regions of a facial image, retraining the model, and then observing the relative importance of each region on the model’s performance^24^. All analyses were performed on a Linux (Ubuntu 22.04.3 LTS) server equipped with an NVIDIA RTX 3060 Ti graphical processing unit and 128 gigabytes of memory.

## Results

Incremental training of the subgroup of 18 White females with vEDS compared to matched CFD controls revealed that strong classification performance could be achieved with as few as 13 images (Fig. 1B), where the model demonstrated an average Brier score of 0.13, an AUC of 0.94 (95% CI 0.8894, 0.9899), and a p-value of 1.35×10^-6^, indicating excellent predictive accuracy with a limited number of training images (Fig. 2B). When applied to the full set of 30 vEDS images, the AI classifier maintained high performance in distinguishing vEDS from an ethnically, age-, and sex-diverse set of controls. It achieved an average Brier score of 0.05, an AUC of 0.99 (95% CI 0.9864, 0.9903) and a p-value of 2.93×10^-11^ (Fig. 2A), confirming the model’s robustness across the broader dataset. An analysis of facial phenotypes associated with the 20 *COL3A1* glycine substitution cases versus the ten non-glycine cases (e.g., in-frame splice site and HI) showed no significant difference between these groups, reflected in a Brier score of 0.29, an AUC of 0.33 (95% CI 0.2683, 0.3937), and a p-value of 0.673 (Fig. 2B).

**Figure 2.**
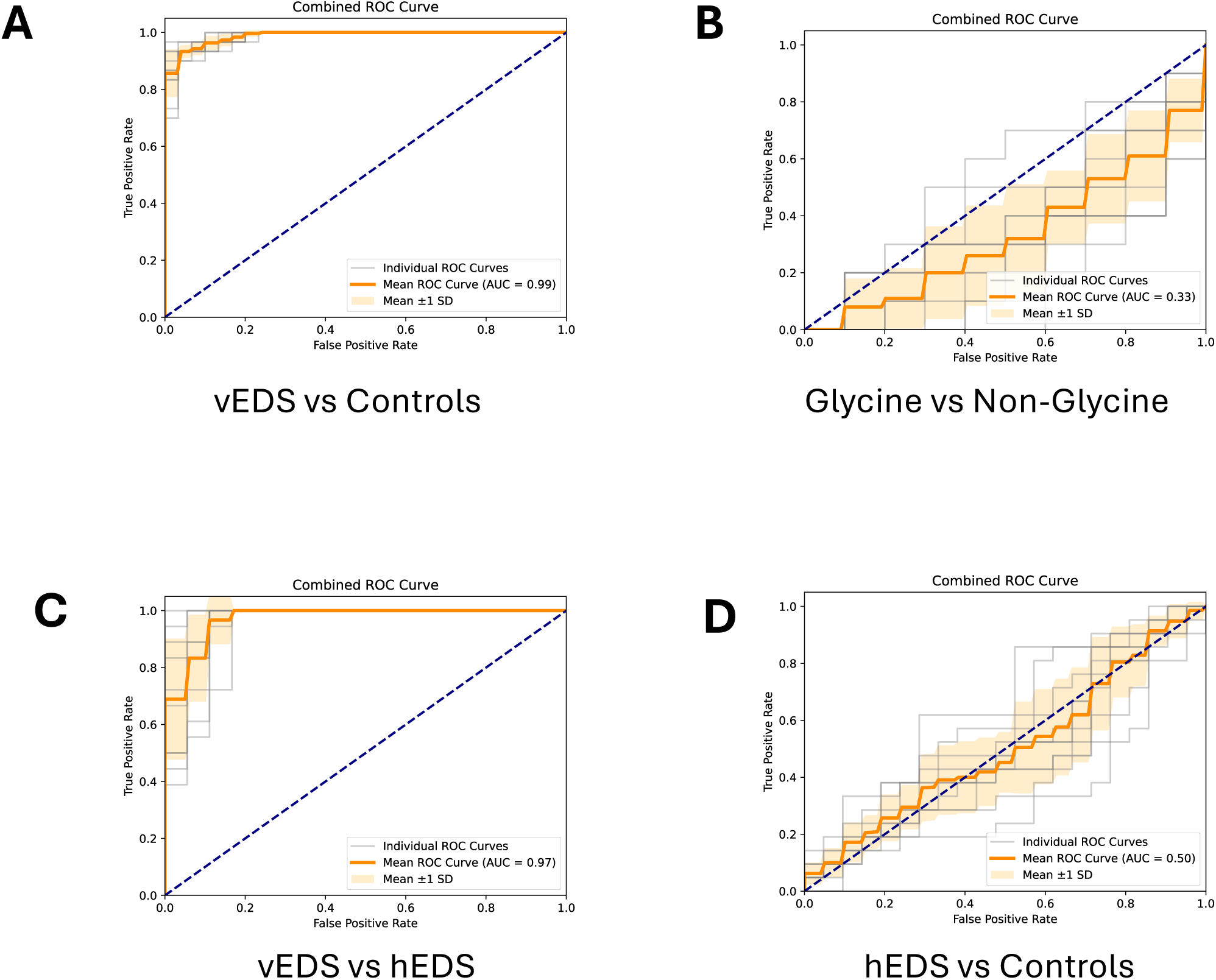
Combined Receiver Operating Characteristic Curves. Curves illustrate the performance of the classifier for different comparisons: vEDS vs. Controls (Panel A), glycine vs. non-glycine *COL3A1* variants (Panel B), vEDS vs. hEDS (Panel C), and hEDS vs. Controls (Panel D). Gray lines represent the individual ROC curves for each of the ten model training iterations. The dark orange line indicates the mean ROC curve, with the shaded region depicting the standard deviation (±1 SD) around the mean. The mean ROC AUC is reported as a measure of the classifier’s overall discriminative performance. ROC, Receiver Operating Characteristic; AUC, Area Under the Curve; vEDS, vascular Ehlers-Danlos syndrome; hEDS, hypermobile EDS

Additionally, the model effectively differentiated vEDS from hEDS, achieving an average Brier score of 0.09, AUC of 0.97 (95% CI 0.9553, 0.9878), and a p-value of 1.12×10^−19^ (Fig. 2C). This high level of accuracy indicates the model’s ability to distinguish facial features unique to vEDS that are not present in another more common form of EDS, hEDS. Finally, LIME-generated heatmaps showed that the model prioritized areas of the face containing known characteristic facial features seen in vEDS, such as the eyes, nose, and lips^24^ (Fig. 3A-D). This alignment with clinical descriptions supports the model’s validity in capturing vEDS-specific facial features that are recognized by experienced geneticists. No statistically significant difference was observed between the hEDS facial images and matched control groups, with a Brier score of 0.25, an AUC of 0.50 (95% CI 0.4488, 0.5580), and a p-value of 0.25 (Fig. 2D). These results suggest that individuals with hEDS have facial features similar to those of the general population.

**Figure 3:**
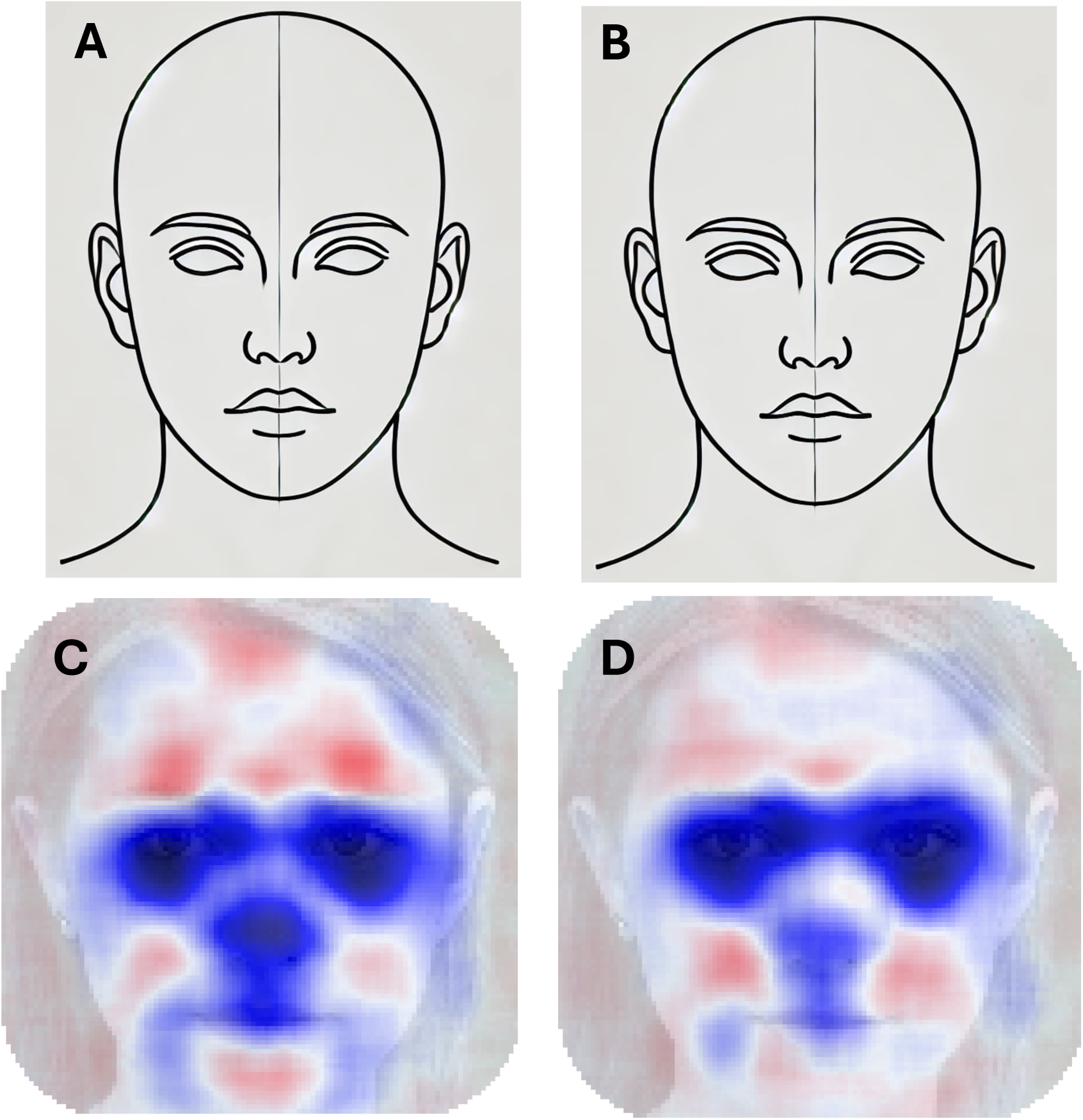
Spectrum of vEDS-associated Facial Features. Panel A shows an individual* with vEDS featuring the characteristic narrow nose, prominent eyes, and a thin vermilion of the lips typical for pathogenic *COL3A1* glycine substitutions. Panel B depicts an individual* with vEDS due to a pathogenic *COL3A1* HI variant but with a milder facial phenotype. LIME heatmaps indicating the facial areas most important to the model for identifying vEDS and distinguishing it from controls (Panel C) and hEDS (Panel D) are shown. The heatmap backgrounds use a StyleGAN-generated synthetic face^33^. Areas highlighted in blue, such as the eyes, nose, and lips, correlate with the vEDS cases and align with the known characteristic facial appearance of vEDS. Conversely, red areas correlate with control or hEDS cases. The intensity of the blue and red colors reflects the model’s confidence, where deeper hues indicate stronger associations while lighter hues suggest a weaker contribution. vEDS, vascular Ehlers-Danlos syndrome; HI, haploinsufficiency; hEDS, hypermobile EDS; and LIME, Local Interpretable Model-agnostic Explanations

## Discussion

This study demonstrates the potential of an AI-based facial recognition model, trained on a small number of vEDS facial images, to effectively distinguish individuals with vEDS from those in the general population and hEDS cases, achieving an AUC ≥ 0.97 with both groups. The LIME data indicate that the AI model effectively focuses on facial regions containing characteristic vEDS facial features, such as prominent eyes, narrow nose, and thin vermilion of the lips^25^, thus further validating the results.

Ehlers-Danlos syndrome (EDS) is a group of 13 conditions grouped together based on common systemic features, which include joint hypermobility, skin hyperextensibility, and abnormal scar formation^5^. vEDS is the only subtype of EDS associated with a highly penetrant risk for life-threatening complications, i.e., spontaneous arterial and GI ruptures. The most common EDS subtype is hEDS, characterized by joint hypermobility, variably associated with GI issues that do not include spontaneous rupture of the gut but rather chronic pain and autonomic dysfunction^5^. Notably, no genes have been identified for hEDS, making it a clinical diagnosis only. Given that hEDS accounts for 90% of all EDS cases^25^, vEDS needs to be differentiated from hEDS in clinical settings since vEDS is associated with life-threatening vascular and GI complications while hEDS is not^1,2^. Thus, facial images are potentially a rapid and cost-effective way to distinguish hEDS from vEDS.

The study also investigated how different *COL3A1* variant types influence the model’s overall performance, given the strong association between variant type and the severity of the vascular phenotype. For example, missense variants that disrupt glycine residues in the triple helical domain of type III collagen, as well as splice-site variants leading to in-frame exon skipping, are associated with earlier onset and more severe complications, as well as more pronounced facial features^26,27^, when compared to haploinsufficiency (HI) variants^3,27^. Although the analysis was underpowered due to the limited number of splice-site and very few HI variants, no clear difference in facial features was observed between glycine and non-glycine variants. This could suggest that the model may be capable of recognizing vEDS across the variant spectrum. However, a larger dataset is needed to confirm this hypothesis and enable more granular analyses of different variant types.

This AI-based method presents a valuable tool in resource-limited areas, where access to genetic testing is limited but medical therapies recommended for vEDS to prevent arterial events are accessible. For example, beta-blockers have been shown to reduce vascular events in patients with vEDS^28^, prompting the initiation of a clinical trial currently underway (ClinicalTrials.gov Identifier: NCT05432466). Additionally, using facial images to diagnose vEDS would be useful for pediatricians, who may see many children with loose joints and can apply this method to identify the rare vEDS case, and emergency physicians, who need to make rapid diagnoses for correct management. By reducing dependence on genetic testing, which can be expensive and less accessible in resource-limited areas, AI-based facial analysis could democratize the early diagnosis of vEDS, allowing for optimal management and early recognition of adverse events, and therefore improve outcomes globally.

There are several limitations to this study, primarily related to the relatively small sample size and limited ethnic diversity. Facial recognition models generally perform better with larger datasets^29^, which presents a challenge given the rarity of vEDS, with an estimated prevalence of 1 in 50,000 to 1 in 200,000 individuals^1^. Additionally, studies have shown that model performance can vary based on the ethnicity of the individuals used for training^30,31^, underscoring the importance of diversity in such datasets. To address these limitations, we first applied the principle of transfer learning by using the VGGFace2 pre-trained model, which was trained on an ethnically, age, and gender-diverse dataset of over 3 million facial images^21^. This approach allowed us to extract facial features from our data and subsequently train the SVM on a smaller vEDS dataset while maintaining strong performance and reducing the risk of overfitting. We also utilized the Chicago Face Database^19^, a similarly diverse control dataset, to represent the general unaffected population, further preventing overfitting. These strategies collectively enabled us to achieve strong discriminatory performance with as few as thirteen images. Nevertheless, expanding the dataset to include a broader range of ethnic groups, age ranges, sexes, and variant types would likely enhance the model’s robustness and overall generalizability. Lastly, given the privacy concerns associated with facial images, we could explore using alternative image types, including of the hand, where individuals with vEDS are known to exhibit an aged, acrogeric appearance^5,32^.

Ideally, this tool could be developed into a mobile application for healthcare providers. As a screening tool, its primary goal would be to detect as many cases as possible, given that early diagnosis is critical. Additionally, it could help assess the significance of *COL3A1* variants of uncertain significance (VUS) identified on genetic testing by indicating whether an individual has the characteristic vEDS facial features. However, because vEDS is a rare condition, it’s equally important to minimize false positives to avoid overwhelming healthcare systems and causing undue anxiety. Currently, the tool provides a binary output indicating whether an individual is likely to have vEDS based on the SVM model’s calculated probability. To optimize both sensitivity and specificity, it will be essential to establish an ideal threshold that balances the need for accurate detection with the risk of false positives. Integrating this tool into clinical practice will also require ensuring data privacy, managing the potential for algorithmic bias, and making AI technology widely accessible to healthcare providers.

In conclusion, this study highlights the potential of AI-based facial recognition as a screening tool for vEDS, offering a non-invasive, cost-effective method to aid early diagnosis and improve patient outcomes. By providing a more precise and timely identification of vEDS, such approaches can help implement guideline-based management strategies that could significantly reduce the risk of severe vascular complications in affected individuals. Equally important is it has the potential to be used as a method to exclude vEDS in patients with overlapping skin and joint complications but who lack the risk for vascular and GI complications. Further research, along with careful consideration of ethical and practical aspects, will be crucial to enhancing the model’s capabilities and ensuring its successful integration into routine clinical practice, particularly in resource-limited settings.

## Acknowledgements

We extend our appreciation to individuals and families who participated in this study and collaborated with us on this report.

## Data Availability

Study participant images are not shareable due to IRB restrictions. Trained models will be made available on GitHub.

## CRediT Author Statement

**David R. Murdock:** Conceptualization, Methodology, Software, Formal analysis, Validation, Investigation, Writing - Original Draft. **Adarsh Suresh**: Methodology, Writing - Original Draft. **Ernesto Calderon Martinez:** Resources, Data Curation, Writing - Review & Editing. **Isabella Marin**: Methodology, Resources, Writing - Review & Editing. **Frances Marin**: Methodology, Resources, Writing - Review & Editing. **Alan C. Braverman**: Methodology, Resources, Writing - Review & Editing. **Angela T. Yetman**: Methodology, Resources, Writing - Review & Editing. **Shaine A. Morris**: Methodology, Resources, Writing - Review & Editing. **Dianna M. Milewicz:** Conceptualization, Methodology, Resources, Supervision, Funding acquisition, Writing - Review & Editing.

## Ethics Declaration

This study was reviewed and approved by the Institutional Review Boards of UTHealth and the sites of recruitment. Signed informed consent and/or authorization to use facial image data for research and publication were obtained by participating institutions and are on file.

*Placeholder images, please contact authors to obtain these materials.

